# Prospective validation of a predictive non-invasive SNP-based biomarker signature in resectable pancreatic cancer: a study protocol

**DOI:** 10.1101/2023.10.24.23297456

**Authors:** Nico Seeger, Stefan Gutknecht, Irin Zschokke, Isabella Fleischmann, Nadja Roth, Juerg Metzger, Markus Weber, Stefan Breitenstein, Lukasz Filip Grochola

## Abstract

**Background:** A growing body of work suggests that inherited genetic variants can be utilized as non-invasive biomarkers to predict cancer-specific patient outcomes and guide therapy after resection of pancreatic ductal adenocarcinoma (PDAC). Specifically, two recent retrospective analyses led to the identification and retrospective validation of three single nucleotide polymorphisms (SNPs) in the CD44 and CHI3L2 genes (SNPrs187115, SNPrs353630, SNPrs684559) genes that can serve as predictive biomarkers to help select patients who are likely to benefit from pancreatic resection. Those SNPs have been shown to associate with an over 2-fold increased risk for tumour-related death in three independent PDAC study cohorts from Europe and the U.S (Hazard ratio (HR) up-to 0.38; p-value up-to 1x10^−8^).

**Methods:** In order to translate these retrospective findings into clinical practice, we aim to utilise a cohort of PDAC patients who undergo pancreatic resection in three hospitals in Switzerland to prospectively validate the association of the identified SNPs in the CD44 and CHI3L2 genes with PDAC survival after resection in a controlled clinical setting. All patients with a histopathological diagnosis of PDAC who undergo pancreatic resection and fulfil the inclusion criteria will be included consecutively. The SNP genotypes will be determined using standard genotyping techniques from patient blood samples. For each genotyped locus, log-rank and Cox multivariate regression tests will be performed, accounting for the relevant covariates AJCC-stage and resection-status. Clinical follow-up data will be collected for at least 3 years. Sample size calculation resulted in a number of 150 patients (80% power to detect allelic differences in survival; effect size>2.0 between genotypes; two-sided α-level of 0.05; drop-out rate of 10%; potential additional error margin of 10%).

**Discussion:** This is the first prospective study of the CD44 and CHI3L2 gene SNP-based biomarker signature in PDAC. A prospective validation of these biomarkers would enable its utilization as a non-invasive, predictive signature of survival after pancreatic resection that is readily available at the time of PDAC diagnosis. The results of this study may lead to improved patient outcomes in resectable PDAC.

**Trial registration:** Not listed in a registry, because no results of a health care intervention are reported.

## Background

Pancreatic ductal adenocarcinoma (PDAC) is the most frequent type of pancreatic cancer and one of the most lethal malignancies due to an early local invasion and high metastatic potential (1, 2). In spite of this aggressive phenotype, it shows a wide range of biology, whereby early recurrence is observed in some patients who undergo pancreatic resection and long-term cancer-free survival can be achieved in others (3-5). It is thought that such individual oncologic courses and the response to personalized treatment decisions can be predicted by the understanding of human germline genetic variation (6, 7). Importantly, this genetic information can be found in the germline DNA and does neither require a tumour biopsy nor the technically demanding isolation of circulating tumour cells. Instead, it can be easily obtained by a simple and cheap blood or saliva test at any time during diagnostic workup or treatment and can therefore be utilized as a non-invasive predictive biomarker at the time of diagnosis. Indeed, this type of personalized medicine is no longer a fantasy but a maturing reality. In other cancer types, such as acute lymphoblastic leukaemia, colorectal, lung and breast cancer, the genotyping of certain single nucleotide polymorphisms (SNPs) is recommended by the US Food and Drug Administration (FDA) (8).

In pancreatic cancer, a growing body of work suggests that inherited genetic variants can also be utilised to predict cancer-specific patient outcomes and guide therapy. Specifically, in a retrospective analysis, we identified a germline variant in the CD44 gene (SNPrs187115) that can serve as a predictive biomarker to help select patients who are likely to benefit from pancreatic resection (9, 10). More recently, we performed a genome-wide screen for inherited variants that affect survival of PDAC based on existing biological knowledge about the regions where the polymorphisms reside (11). This search for high-frequency polymorphic variants that affect tumour-related survival and either result in an altered protein structure and function or reside in known regulatory, non-coding genomic regions has identified two clinically-relevant polymorphisms in functional regions of genes which are known to regulate cancer progression, invasion and metastasis(11). Specifically, those regulatory variants within the CHI3L2 (SNPrs684559) and CD44 genes (SNPrs353630) were shown to associate with an over 2-fold increased risk for tumour-related death in two independent retrospective PDAC study cohorts from Europe and the U.S (Hazard ratio (HR)=0.38; 95% Confidence Interval (CI)=0.27-0.53; p-value=1x10^−8^, q-value<0.05) (11).

Taken together, those retrospective studies have demonstrated the potential of polymorphic variants to identify a subset of high-risk pancreatic cancer patients with very low survival probability that might be eligible for inclusion in clinical trials of new therapeutic strategies, including neoadjuvant chemotherapy protocols and novel adjuvant systemic regimens. In addition, they suggest that the biological knowledge about these SNPs could help guide the development of such individualized genomic treatment strategies.

However, despite their robustness, the described results have not yet been validated prospectively and it is unclear whether those observations can be independently observed in a controlled clinical setting.

## Methods/Design

### Aim, design and setting of the study

The project is designed as a one-arm prospective controlled clinical study which aims to validate the association of the previously identified SNPs in the CD44 and CHI3L2 genes with PDAC survival after tumour resection. We aim to utilise a prospective cohort of PDAC patients who will undergo pancreatic resection at our established network of three cantonal hospitals (Departments of Surgery at the Cantonal Hospital of Winterthur, Cantonal Hospital of Lucerne and the Triemli Hospital in Zurich) in Switzerland.

#### Characteristics of participants

Patients with a diagnosis of a pancreatic tumour mass, either biopsy-proven or highly indicative of PDAC will be recruited before surgery in the outpatient clinic at the participating centres after completed standard pre-operative evaluation of pancreatic cancer diagnosis and assessment of tumour resectability according to the best standard of care and the following criteria:

1. Pancreatic tumour mass either biopsy-proven or highly indicative (as assessed by pre-operative MRI and/or CT and/or EUS imaging) of PDAC in the head or tail of the pancreas
2. TNM Stage: c/uT1-4, c/uNx, cM0 (AJCC 7e)
3. Open or minimally-invasive pancreatic resection, including conversion laparotomy
4. Patients who undergo primary resection of their tumours
5. Patients after neoadjuvant chemotherapy and/or radio-/chemotherapy
6. Patients who previously underwent pancreatic resection for reasons other than *ductal adenocarcinoma of the pancreas*
7. Age ≥ 18 years
8. Written informed consent.

Patients who subsequently successfully underwent tumour resection and whose diagnoses of a *ductal adenocarcinoma of the pancreas* has been confirmed by histopathological examination of the resected specimen will be included in the study. All patients who fulfil those criteria will be included, regardless of the tumor resection margins (R0, R1, R2) and regardless of intra- or postoperative mortality.

The presence of one of the following criteria will lead to the exclusion of the subject:

1. Any histopathological entity other than *ductal adenocarcinoma of the pancreas*, including but not limited to:
  a. Adenocarcinoma of the papilla of Vater
  b. Acinar cell carcinoma
  c. Distal cholangiocarcinoma
  d. Neuroendocrine tumours including neuroendocrine carcinoma
2. Metastatic disease, including intraoperative findings confirmed by histopathological examination (cM1, pM1)
3. Patients who had previously undergone pancreatic resection for *ductal adenocarcinoma of the pancreas* and present either with:
  a. Local recurrence or
  b. Second primary in the pancreatic remnant and had not been included in the study yet. However, individuals who had already been included at the time of the first resection of a *ductal adenocarcinoma of the pancreas* will not be excluded from the study retrospectively because of a local recurrence or a second primary but will continue to receive a follow-up.
4. Age < 18 years
5. No written informed consent.

All patients that have been excluded from participation will be recorded together with the reason for the exclusion.

### Endpoints and data collection

The primary endpoint of the study is the tumour-related survival. Secondary endpoints include recurrence- and overall survival. The exact type and time of data collection is listed in detail in Table 1. All data relevant to this study will be coded (biological material and health-related personal data), safely protected and recorded in a digital format using a GCP-compliant EDC system. The estimated total duration of the study is 5 years, including the scheduled 3-year postoperative follow-up.

**Table 1.**
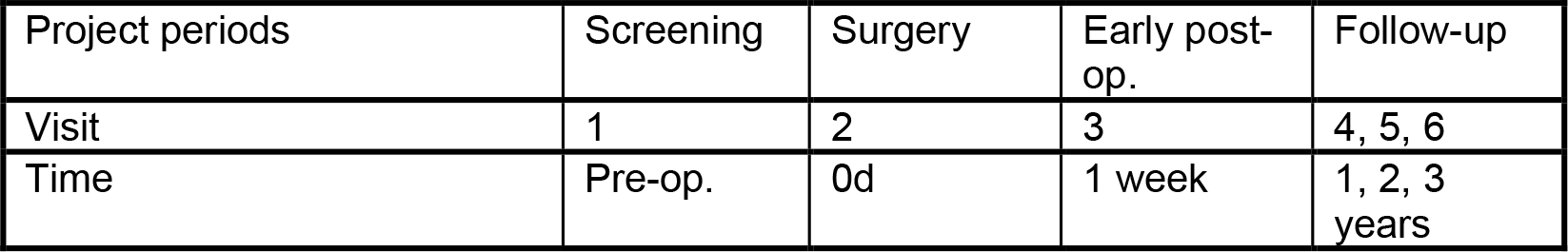
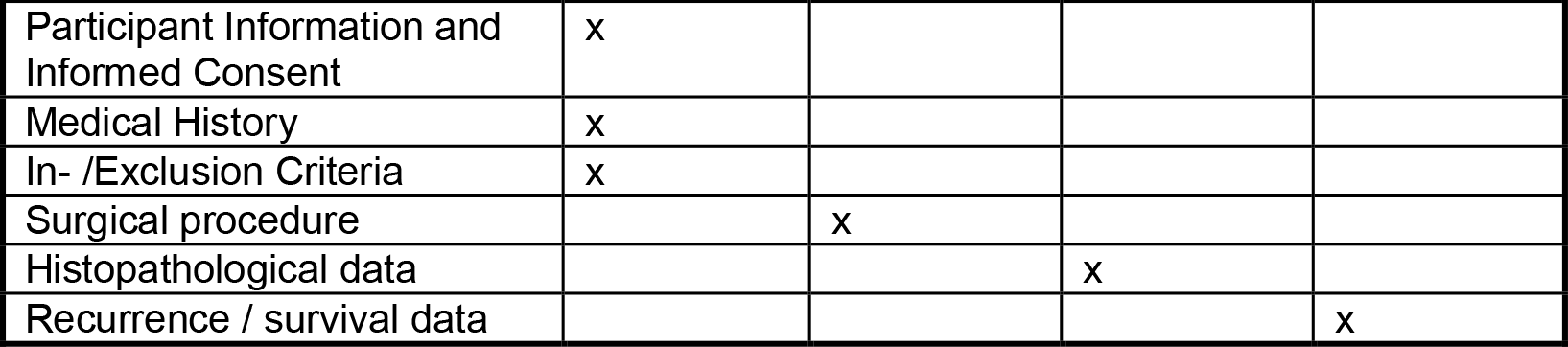

All cases will be presented at an interdisciplinary tumour-board meetings at each participating centre pre- and postoperatively. All patients will be offered standard of care multimodal treatment at the discretion of the interdisciplinary tumour board at each participating centre. The operation will be performed according to the group assignment by senior surgeons only, who have extensive experience in pancreatic resections. All surgeries will be performed at the Departments of Surgery at the Cantonal Hospital of Winterthur, Cantonal Hospital of Lucerne and the Triemli Hospital in Zurich. The follow-up of the patient will be recorded for at least 3 years after tumour resection. It will be completed using the hospital’s own records or by contacting the patient’s general practitioner or oncologist and therefore no extra appointments in the outpatient clinic are necessary for study purposes.

### Power of the study

Sample-size calculations are based on the results of the previously mentioned retrospective analyses with a hazard ratio (HR) of approximately 2.0 between genotypes (9-11). This study is designed to detect a hazard ratio (HR) of > 2.0 between genotypes. With the use of a two-sided α level of 0.05, and considering a drop-out rate of 10% as well as a potential additional error margin of 10%, 150 patients are required to obtain 80% power to detect a difference in cancer-related survival (G-Power 3.1 software, Heinrich-Heine University, Germany)

### Statistical analysis

A parametric two-sided Student’s T-Test and a two-tailed Mann-Whitney test will be used to compare data distributions in groups with normally distributed data and data that deviate from normal distribution, respectively. Fisher’s Exact test will be used to analyse frequency distributions in cross-tables. Kaplan-Meier survival curves (log-rank test) and Cox proportional hazards models will be calculated to assess survival times. All statistical analyses will be performed using the software R and SPSS.

### Ethics

The study will be carried out in accordance with principles enunciated in the current version of the Declaration of Helsinki, the guidelines of Good Clinical Practice (GCP) issued by ICH, and Swiss regulatory authority’s requirements. The study has been approved by the local ethics committee (Kantonale Ethikkommission Zürich, BASEC-Nr. 2019-00178).

## Discussion

This is the first prospective study of the CD44 and CHI3L2 gene SNP-based biomarkers in ductal adenocarcinoma of the pancreas after tumour resection. A prospective validation of these biomarkers would allow its utilization as a predictive signature of survival after pancreatic resection that is readily available at the time of PDAC diagnosis. The genotype of the identified SNPs can be determined before the start of any treatment by a technically simple blood test and utilised to identify patients who are at higher risk for faster tumor progression. Other prognostic factors currently in use or suggested to be useful in a clinical setting such as completeness of resection, lymph-node involvement or postoperative carbohydrate antigen 19-9 (CA19-9) serum levels can only be precisely determined after surgical resection and therefore have limited relevance to treatment decisions (12). In contrast, SNP-based biomarkers can guide preoperative treatment decisions, such as the decision to perform neoadjuvant chemotherapy in patients with an aggressive tumor biology or affect adjuvant treatment protocols. The results of this prospective study may lead to improved patient outcomes in patients with pancreatic ductal adenocarcinoma that is amenable to resection.

## Data Availability

All data produced in the present study are available upon reasonable request to the authors

## Declarations

### Ethics approval and consent to participate

Ethics approval: The study has been approved by the local ethics committee (Kantonale Ethikkommission Zürich, BASEC-Nr. 2019-00178).

#### Consent for publication

Not applicable

### Availability of data and materials

The datasets used and/or analysed during the current study are available from the corresponding author on reasonable request.

### Authors’ contributions

NS and LFG designed the protocol and drafted the manuscript. All other authors participated in the design of the study and are local investigators at the Cantonal Hospital of Winterthur, Cantonal Hospital of Lucerne and the Triemli Hospital in Zurich. All authors were involved in editing the manuscript and approved the final text of the manuscript.

## Acknowledgements

None.

